# Caregivers’ and nurses’ perceptions of the Smart Discharges Program for children with sepsis in Uganda: A descriptive qualitative study

**DOI:** 10.1101/2023.07.18.23292842

**Authors:** Justine Behan, Olive Kabajaasi, Brooklyn Derksen, George Sendegye, Brenda Kugumikiriza, Clare Komugisha, Radhika Sundararajan, Shevin T. Jacob, Nathan Kenya-Mugisha, Matthew O. Wiens

## Abstract

Sepsis arises when the body’s response to infection results in organ dysfunction. Among children hospitalized with suspected sepsis in low-income country settings, mortality rates following discharge are similar to mortality rates in hospital. The Smart Discharges Program uses a mobile health (mHealth) platform to identify children at high risk of post-discharge mortality to receive enhanced post-discharge care. This study sought to explore the perceptions of the caregivers and nurses of children enrolled into the Smart Discharges Program. We conducted a descriptive qualitative study that used a phenomenological approach. We conducted in-person focus group discussions (FGDs) with 30 caregivers of pediatric patients enrolled in the Smart Discharges Program and individual, semi-structured interviews with eight Smart Discharges Program nurses. The study was carried out at four hospitals in Uganda in 2019.

Following thematic analysis, three key themes pertaining to the Smart Discharges program were identified: (1) Facilitators and barriers to follow-up care after discharge; (2) Changed behavior following discharge; and (3) Increased involvement of male caregivers. Facilitators included telephone/text message reminders, positive nurse-patient relationship, and the complementary aspects of the program. Resource constraints and negative experiences during post-discharge care seeking were reported as the most prominent barriers to post-discharge follow up. When provided with relevant and well-timed information, caregivers reported increased knowledge about post-discharge care and improvements in their ability to care for their child. Enrolment in the Smart Discharges Program also increased male caregiver involvement, which was reported as improved engagement in care, increased provision of resources and improved communication within the family and with the healthcare system. The Smart Discharges approach is an impactful strategy to improve pediatric post-discharge care, and similar approaches should be considered to improve the hospital to home transition in similar low-income country settings.

## Introduction

Sepsis arises when the body’s response to an infection results in organ dysfunction (1). In 2017, nearly 49 million incident cases of sepsis were reported globally with half occurring in children under five (2). The global sepsis burden is disproportionately carried by low-and middle-income countries (LMICs), with close to 3 million deaths annually among children under the age of five, most of which occur in sub-Saharan Africa (2).

Among children hospitalized with suspected sepsis in low-income country settings, mortality rates following hospital discharge are similar to those seen during the hospital phase of illness (3-8). Often referred to as post-sepsis syndrome, this period of high vulnerability following discharge is often unknown to many parents and healthcare workers who are also poorly equipped to facilitate a robust recovery within a community environment (9, 10). Recent data suggest that improved post-discharge outcomes can be achieved through educational interventions and community-level referrals for follow-up (11). Such interventions may form critical components necessary to improve the long-term survival of children with sepsis (12). However, despite their potential value, financial barriers faced by families and children, such as transportation costs, facility fees, and medications prescribed upon discharge, may make the implementation of such programs difficult (5, 13).

The Smart Discharges Program uses a mobile health (mHealth) platform to identify children at high risk of post-discharge mortality for enhanced post-discharge care. This innovative program provides an opportunity for front-line health workers in resource-poor environments to identify vulnerable children and to streamline a path for follow-up care during the post-discharge period (14). Counselling and discharge planning target factors that are known or assumed to contribute to post-discharge mortality, including clinical, social, and system considerations. While all participants receive enhanced discharge counselling, high-risk children receive down-referrals to community health facilities for follow-up (12).

The Smart Discharge Program is currently being evaluated to determine its impact on post-discharge mortality among children admitted with suspected sepsis in Uganda (15). However, the perceptions and experiences of the parents/caregivers and nurses participating in this program have not yet been explored. The caregivers and nurses experiences are important to provide valuable insights applicable to program improvement. Therefore, we designed a qualitative study to explore the parents’/caregivers’ and nurses’ perceptions and experiences of the Smart Discharges Program and its effects on the post-discharge care of children who have been hospitalized with suspected sepsis.

## Methodology

### Design

We conducted a qualitative study using a descriptive phenomenological approach (16). This approach was well suited for our study because the goal is understanding the lived experiences of participants (17). Data were collected through in-person focus group discussions (FGDs) with caregivers of pediatric patients enrolled in the Smart Discharges Program and individual, semi-structured interviews with Smart Discharges Program nurses who had provided counselling to the caregivers during the child’s admission. The study is reported using the Consolidated Criteria for Reporting Qualitative Research (COREQ) (Supplementary File S1) (18).

### Study setting

The study was carried out at four hospitals in Uganda, including three public regional referral hospitals (RRHs) and one private-not for profit hospital (Table 1). Caregivers included in the study were from the surrounding areas, but their children received care and were enrolled in the Smart Discharges Program at these four facilities.

**Table 1:**
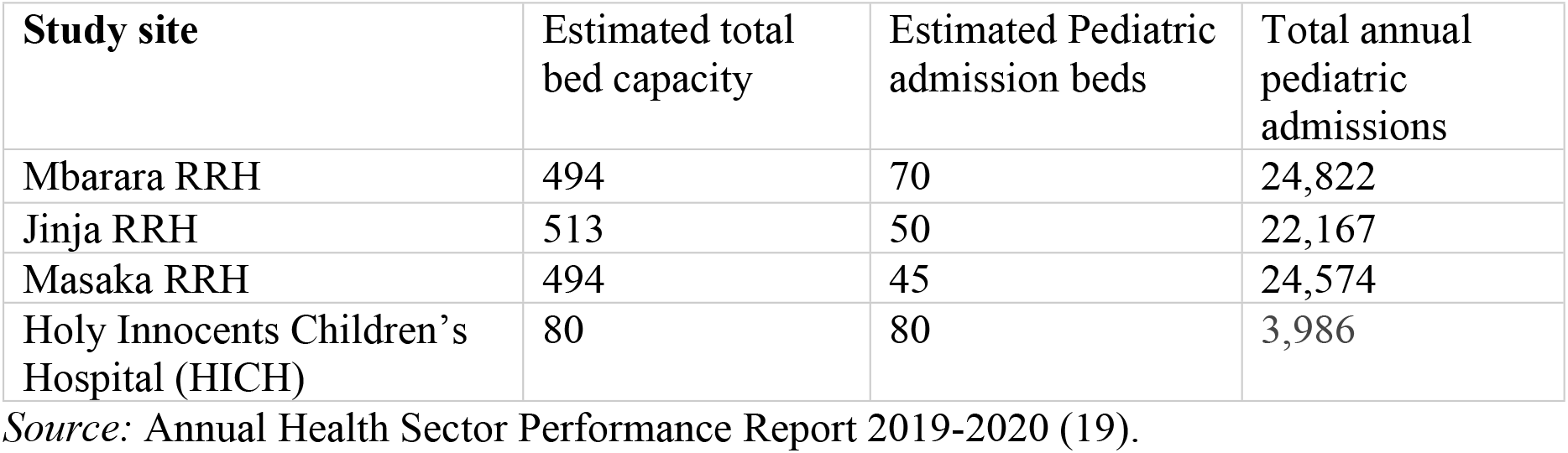
Annual Hospital Pediatric Admissions for 2019/2020.

### Sample selection, recruitment, and consent

Participants were purposively selected for the caregiver FGDs with primary consideration given to ensuring balance between key post-discharge outcomes, including readmission, post-discharge mortality as well as adherence and non-adherence to prescribed post-discharge follow-up. Caregivers were eligible if their child had been enrolled into the Smart Discharges program and had been discharged at least two months prior to recruitment. Nurses were eligible to participate in the in-depth interview if they had worked in the Smart Discharges program for at least two months. FGD participants and nurses were invited to participate through telephone calls from the social scientist (OK), and FGDs and interviews were conducted in person in private rooms within the pediatric wards at the study sites. All participants provided written informed consent.

### Data collection

FGDs and in-depth interviews were conducted between August 29, 2019 and October 30, 2019. Three female and one male local research assistants (including co-authors GS, BK, CK) who were fluent in the languages (Runyankore, Luganda, and Lusoga) and had 2-4 years experience in moderating FGDs conducted the FGDs. They were trained and supervised by a female social scientist (OK; MA Sociology), with relevant working experience in conducting qualitative research in Ugandan health settings. Facilitators of the same sex as the participants moderated the FGDs in the local language. Individual interviews with nurses were conducted by OK in English. A focus group discussion guide (Supplementary File S2) and a semi-structured interview guide (Supplementary File S3) were used to provide structure and consistency to the discussion/interviews while allowing for novel concepts to be shared. The FGD guide focused on description of the experience at the admitting facility, experience after discharge, referral for follow-up, referral completion, referral facilities, and barriers to providing adequate care. Data collection was considered complete following four FGDs, based on the richness of data shared through feedback from the moderators, review of the transcripts, and notes. The in-depth interviews focused on the experience of providing caregiver education and counselling, and reporting of which programmatic components worked well and did not work well. Nurse interviewees were also asked about their observations of the benefits and challenges of the program to the caregivers. The FGD and interview guides were not pre-tested prior to use and no repeat interviews were done.

### Data analysis

At the end of each FGD, field notes were written and shared with the study Principal Investigator to review and provide feedback to refine the guides for future use. FGDs and interviews were audio recorded and transcribed directly in English by a professional translator and then OK reviewed for accuracy and consistency. Two investigators (JB and OK) independently created the initial coding framework, deductively using the FGDs/interview guide topics and inductively to identify emergent themes and subthemes. The coding framework was refined through discussion between JB, OK, and MOW. During analysis, data was organized using NVivo version 12.0 (QSR, Massachusetts, United States). After development of the coding framework and initial coding, themes were proposed, and discussed between JB, OK, and MOW who jointly agreed on the study themes and then confirmed full team agreement on the final themes.

### Trustworthiness

Trustworthiness and phenomenological validity were achieved through credibility, reflexivity, dependability, conformability, and transferability (17, 20). Themes and sub-themes were triangulated against the field notes of the research assistant who completed the FGDs to ensure credibility of findings during the analytical process. Transcripts were not returned to participants for review and correction. To ensure reflexivity, two researchers (JB and OK) analyzed the data and discussed their findings to produce the coding framework. They discussed potential biases, questions, and concerns, allowing for a sharing of perspectives (21). The research team ensured dependability and confirmability by writing detailed notes and conducting regular discussions to review progress, the research process, and study findings. To improve the transferability and relevance of the results, we provided contextual information about the study setting, sample characteristics, and interview guides.

### Ethical consideration

The Makerere University School of Public Health, Research and Ethics Committee (SPH- REC # 691) and the Uganda National Council for Science and Technology (UNCST SS #5047) provided ethics approval for this study. Participant confidentiality was emphasized during the consent process, and compensation of 25,000 Ugandan Shillings (approximately 7 USD) was given to each participant.

## Results

A total of 40 participants were invited to take part in the study; of these, 38 participated, and two caregivers declined citing lack of transport and time (see Table 2a and 2b: Participant Demographics). Four FGDs were conducted, consisting of one with fathers and three with mothers or other female caretakers. Eight interviews with nurses were completed, two from each participating facility. Each FGD consisted of 7-8 participants and lasted between 90 and 120 minutes, while individual interviews lasted between 45 and 60 minutes.

**Table 2a:**
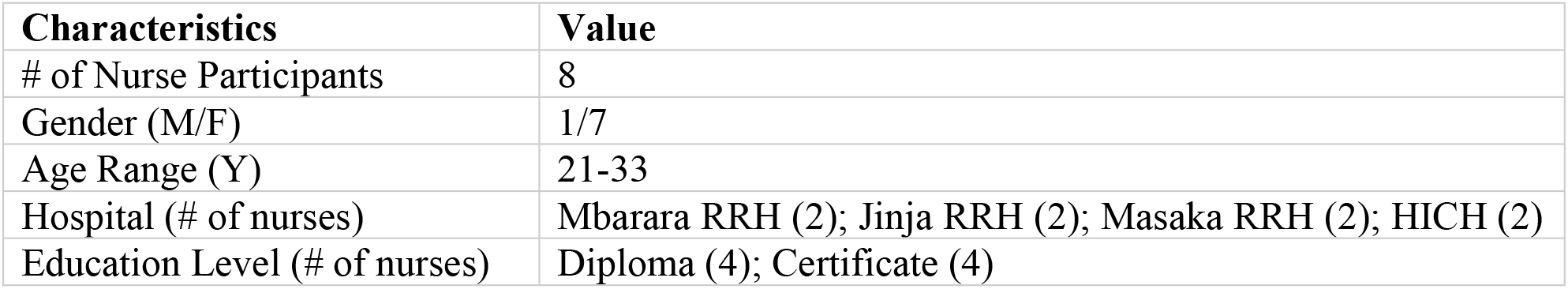
**Nurse Participant Demographics (Individual Interviews)**

| Characteristics | Value |
| --- | --- |
| # of Nurse Participants | 8 |
| Gender (M/F) | 1/7 |
| Age Range (Y) | 21-33 |
| Hospital (# of nurses) | Mbarara RRH (2); Jinja RRH (2); Masaka RRH (2); HICH (2) |
| Education Level (# of nurses) | Diploma (4); Certificate (4) |

**Table 2b:** **Caregiver Participant Demographics (FGDs)**

|  | # of Participants | Age Range (Years) |
| --- | --- | --- |
| Female Caregiver FGD #1 | 8 | 22-45 |
| Female Caregiver FGD #2 | 7 | 28-37 |
| Female Caregiver FGD #3 | 8 | 21-48 |
| Male Caregiver FGD | 7 | 22-57 |
| TOTAL | 30 | 21-57 |

Twenty-seven of the caregivers were parents of the children enrolled in the Smart Discharges Program, and three were other family members (one grandmother and two aunts). Out of the 30 caregivers, reported completing all three follow up appointments, four had children that were readmitted to hospital after their initial discharge, and one had a child that died following discharge.

### Thematic analysis

Following thematic analysis, three key themes were identified: (1) Facilitators and barriers to follow-up care after discharge; (2) Changed behavior following discharge; and (3) Increased involvement of male caregivers. Within each key theme, multiple subthemes were identified and are summarized in Table 3.

**Table 3:**
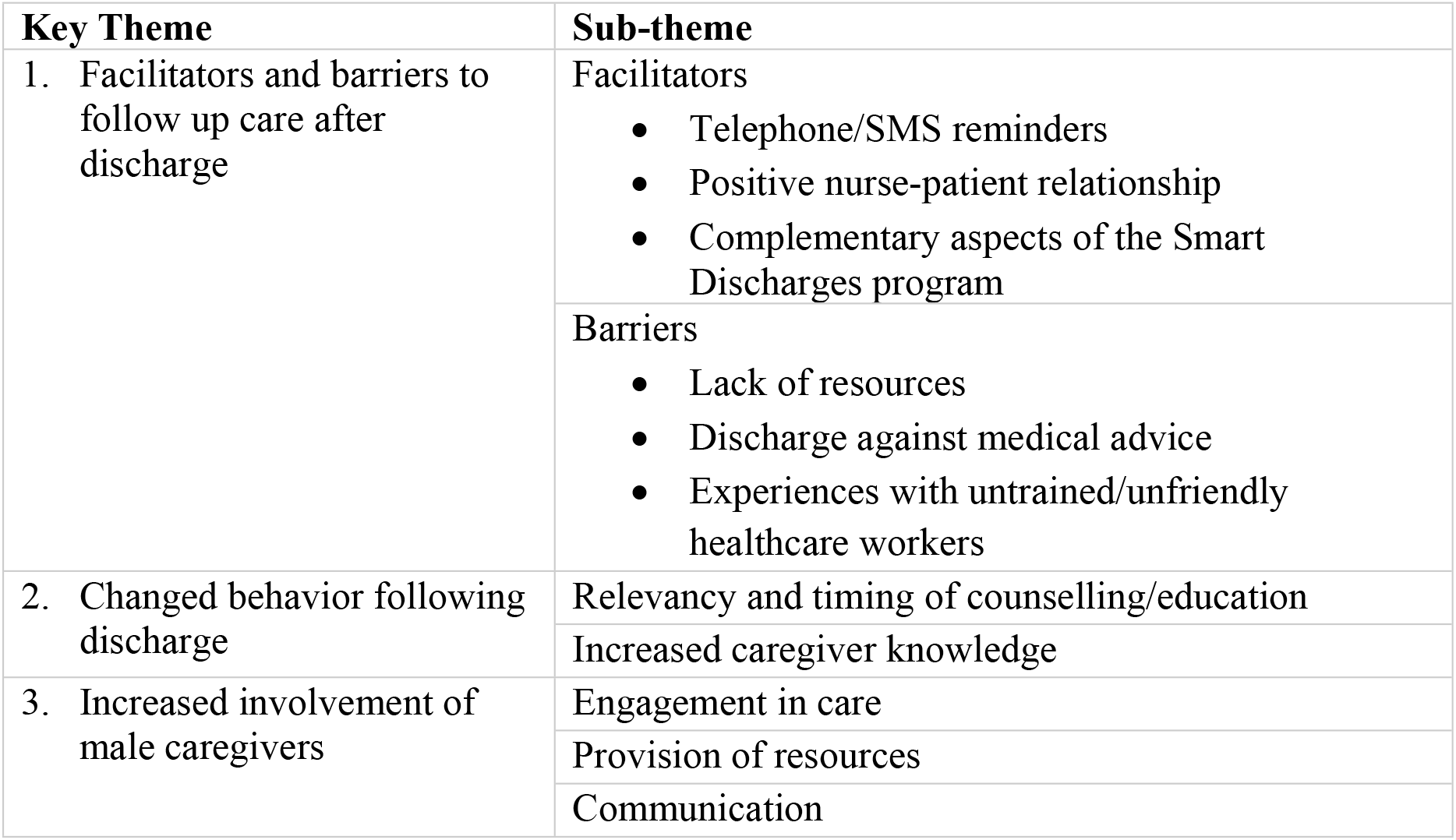
Coding Framework.

### Facilitators and barriers to follow-up care after discharge

#### Facilitators to follow-up care after discharge

The three main facilitators to follow-up care after discharge were telephone/short message service (SMS) text reminders, a positive nurse-caregiver/patient relationship, and the complementary aspects of the Smart Discharges Program. Follow up care following discharge was defined as the adherence to discharge care instructions and attendance at post-discharge follow up visits.

The caregivers were appreciative of and motivated by SMS referral reminders and follow-up phone calls after their discharge. These SMS messages served as a reminder for and provided encouragement to attend the post-discharge follow up at community level facilities.

> “*I finished all the visits because the organization kept on reminding me [via SMS] to take the child to hospital for checkup. This organization of Smart Discharge, they could call me every Tuesday reminding me. [This is] something that encouraged me so I finished all the routines of taking the child to the hospital for checkup*” (Caregiver24).

Nurses reported having more success providing discharge education with caregivers when they had built a positive nursing relationship with the patient and caregiver.

> *“Because it begins with trying to create rapport…and much as the children have come with different infections, when you create that professional relationship they open up… They open up easily and the work becomes easier”* (Nurse3).

Additionally, having a strong nurse-caregiver relationship helped to support better care-seeking behavior after discharge. One nurse shared,

> *“Even after going home, the caregivers keep calling us and telling us how their children are doing and the health centers they have visited. The caregivers now have a positive attitude towards healthcare-seeking than before”* (Nurse1).

Caregivers who were a part of Smart Discharges reported feeling valued and cared for both during the discharge phase of hospitalization as well as after discharge.

Each component of the program, including risk assessment, educational materials, counselling, and post-discharge referral, were described as complementary, and that they “*…work hand in hand”* (Nurse7). A nurse explained, “*[the Smart Discharges Program components] are all complementing each other…they are all supporting one another and none of them can work in isolation”* (Nurse4).

#### Barriers to follow up care after discharge

The key barriers to follow-up care were lack of resources, discharges against medical advice, and experiences with untrained/unfriendly healthcare workers. Lack of resources, which included inadequate financial resources, transportation challenges, and lack of family support, was a prominent barrier to follow-up care. Caregivers’ financial constraints came up frequently and affected follow up visit attendance, adequate nutritional support of recovering children, and purchasing of necessary medication and supplies. One nurse shared,

> *“The high risk patients…When we tell them to go to the nearest health centres some of them are not sure they will have money to do all the three visits…Sometimes they don’t mention this at discharge, but after they have gone home and when we call them they say they did not go for follow up because they did not have money”* (Nurse7).

A caregiver explains this further*, “That day reaches when we don’t even have the money for transport. Even the husband fails to provide money, so you say ‘I will not go that day’”*

(Caregiver10). The nurses highlighted the need to balance the financial demands of having a sick child when there are other children to support at home. It was emphasized that caregivers were doing the best they could with the resources that they have. As another nurse described, *“…the issue of poverty is also a big problem. The caregivers are seemingly appreciating the information being given, but them practicing what is being preached to them is a challenge”* (Nurse1).

Financial constraints also contributed to patients and caregivers being discharged against medical advice. When patients and family leave before it is medically advisable to do so, they miss the opportunity to receive adequate discharge counselling or to receive their follow up appointments. A nurse explained,

> “*There is a financial challenge…For example, some mothers are admitted when they don’t have money to buy food and sometimes they have to buy the drugs as well. So, some will be willing to stay in hospital, but they are worried of what the child will eat. They also see no reason of staying in hospital because they don’t have money to buy the prescribed medicine. So even though we take time to convince the mother to stay, she will eventually go”* (Nurse8).

Negative encounters with healthcare workers at the community health facilities was a barrier identified by nurses as well as caregivers who had attended follow up appointments. These encounters were reported to be negative because not all healthcare workers had received training on the “downward referral” process and, therefore, did not know the process for assessing the children. As a result, healthcare workers were often unfriendly towards patients and caregivers who were reporting for a follow-up assessment. A nurse explained, “*There is a challenge of caregivers who take their children to nearby referral health centres. They face backlashes from health workers there who bash them for taking children to health facilities when they are not sick”* (Nurse1). This was also validated by the caregivers’ perspectives, “*When they were discharging us they told us to take our child to the nearest hospital every week, but when I went there, they shouted at me asking me why am bringing my child to the hospital yet he is not sick”* (Caregiver23). When caregivers had an initial negative experience at a community health facility when seeking follow up care, this often impacted future care seeking and the desire to attend subsequent visits.

### Changed behavior following discharge

The perceptions of both caregivers and nurses demonstrated that the discharge education provided often resulted in changed behavior. Participants noted that appreciating the relevancy and the timing of the counselling was important. When information was relevant and well timed, caregivers’ knowledge about post-discharge care of their children increased, which facilitated behavior changes once they left the hospital.

#### Relevancy and timing of counselling/education

Receptivity of caregivers to discharge education was improved by its perceived relevance and the caregiver’s ability to engage with the materials. As a caregiver explained, *“The education and counseling was useful to me because I got to learn how to take care of a child; how to prepare her meals, hygiene, sleeping under a mosquito net”* (Caregiver2). The education was directly related with how to care for their vulnerable child and relevant to returning home after discharge.

> “*What we do is that we let them know that a time will come and they will be discharged from hospital and what they need to hold as key is that a patient discharged from hospital is not necessarily completely healed but still on the road to recovery*” (Nurse3).

The importance of caregivers receiving education and understanding how it applies to improving outcomes for their children was validated, *“At least if each health worker would talk to a mother and disseminate information regarding the condition of the baby and how to go about it, I am sure this can reduce on the mortalities in their homes”* (Nurse 6). Another key element to receptivity was caregiver engagement with the educational materials and how the educational messages were conveyed. For example,

> “*The [caregivers] are interested in [the educational materials] because it has pictorial illustrations…which caregivers gleefully find illustrative of what they are required to do. For instance, going to hospital, what to feed on, the need for the husband to escort the wife to hospital. At least that one they find it exciting*” (Nurse5).

The timing of education was also found to be important. As a nurse explained,

> “*They don’t know if the child is even getting better, so they tend to be nervous. So, what I do is to find the convenient time when the patient is a bit stable. When there are not many attendants around them, when they are not on phone communicating with their people back home. When they are calm and stable. So, I approach them and talk to them about malaria, what brings about malaria, how it can be prevented at home, and this differs from one child to another depending on how it is presented”* (Nurse3).

By ensuring the caregiver is both physically and mentally in a space where they can learn, they are more able to retain the information.

#### Increased caregiver knowledge

The majority of caregivers shared that at least portions of the education they received from Smart Discharges Program was new information to them. Nurses reported caregivers learning about how to identify danger signs and the critical timing to seek care for their children.

> “*Caregivers are at home but they don’t know about these danger signs. When we talk to them about these, they appreciate and promise to always take their children to nearby health centres as soon as these danger signs manifest so as to get first aid”* (Nurse7).

One father explained a behavior change he observed in his wife,

> “*It wasn’t me they found in the hospital. It was my wife and she learnt many things because I saw her changing. She used not to respond quickly in case of any illness but now she rushes to the hospital at a time she sees a sign of illness*” (Caregiver29).

Many nurses and caregivers spoke about how nutritional education was applied following discharge.

> *“There are some mothers who come with children in hospital. The child is one-year-old but he/she is only feeding on breast milk… there is now some change. Some have been coming here… in our follow up unit… And you find that the child has completely changed in terms of weight addition and generally they look healthy. We attribute this to the information we give them. Because we have been emphasizing to them that the child needs to be introduced to other feeds at 6 months of age, so we feel they have embraced this message. We have also reduced post-discharge mortality and reinfection because of our continued teachings about feeding children on a balanced diet which improves their immune systems to fight against infections”* (Nurse1).

Caregivers also explained that the same principles that were learned through Smart Discharges were applied to the families’ other children. Examples included, feeding all of their children nutritious food and improved care-seeking behaviours when any child in the family fell sick.

A male caregiver shared his experience,

> “*All the things about nutrition, am applying them to my other children and they look healthy. For example, I have chicken, I have fruits like paw-paws* [papaya]*, now I give them, I used to sell all of them and use money to drink. Now I repented”* (Caregiver25).

### Increased involvement of male caregivers

The programs impact on male involvement in their child’s care was seen most prominently through changes in the father’s engagement in care, provision of resources for care, and in changes in health-related communication both within the family and with the healthcare system.

#### Engagement in care

Nurses saw benefits when education/counselling was done with the child’s father present.

> ***“****Some mothers are worried that although they are bound to stay in hospital, it is the husband who should have the final say on this. So in such a situation what I do, I go and give health education to the mother in the presence of the husband and during the session I emphasize the importance of staying in hospital and the husband’s responsibility. I encourage the husband to financially support the mother for the duration of her stay in hospital and in the end both parties agree to stay in hospital”* (Nurse8).

This example highlights how the Smart Discharges education increased male caregivers’ engagement at the facility level. Several caregivers shared examples of how their husbands’ engagement in their children’s care also increased at home.

> “*My husband used not to even give us transport to the hospital when I used to ask him for transport…but since I joined this organization my husband is active. We always come with him to the hospital. He buys food and other responsibilities*” (Caregiver19).

Furthermore, receiving discharge education has given fathers a means to improve their relationships with their children. “*It has helped me in increasing my relationship with my child, because what I read on the card. I buy for her some food like an egg. She becomes so happy*” (Caregiver29).

#### Provision of resources

When the patient’s father received the Smart Discharges education, an increased understanding of the needs of the child and the primary caregiver was seen. This awareness in turn resulted in increased provision of resources for the child and primary caregivers. One caregiver explained,

> “*At the time of discharge, it was not me with the child, it was my husband that was there with the child. He learnt how to care for the child. Since then, when I tell him that I want this for the child, he immediately gives it to me because he does not want to go back to the situation that we were in of spending money in the hospital. Every time I tell him that at least the child has to eat eggs twice a week. He makes sure that he buys the eggs”* (Caregiver21).

Other caregivers shared similar sentiments and acknowledged that after their husbands received discharge education, they bought foods to follow nutritional guidance and purchased the medications prescribed to their children. Male caregivers were more open to providing transportation support for their children to attend follow up appointments or to return to the hospital when the child was sick again. One male caregiver explained, “*When I reached home I made sure that I buy everything he needs [and] take him to the hospital for checkups until he totally healed*” (Caregiver27).

#### Communication

Participation in the program fostered discussions between fathers and mothers about their child’s wellbeing and how to support their vulnerable child. Often the father’s cell phone number was provided to receive follow-up reminders, invoking the father to play an important role in communication with the mother.

> “*Smart Discharge has helped me, in getting involved in the care of my child because whenever I get a call from Smart Discharge, I go and tell my wife that we have to go back to the hospital so that we can learn on how to take care of our child. Therefore, this increases on my relationship with my wife and the child*” (Caregiver28).

Additionally, improved communication between the healthcare system and male caregivers helped them to better understand why their child was hospitalized and to follow discharge instructions.

> “*They are different because previously, they used to just pack for us pills without explaining so much on what you should do. But when we joined Smart Discharge, things changed because before discharging us they first talk to us*” (Caregiver26).

## Discussion

The results of this study improve our understanding of the caregiver and nurse perceptions of a child-centered approach to peri-discharge care, as reflected in the Smart Discharges Program. With the aim of improving outcomes during the vulnerable post-discharge period, this qualitative study found that this program facilitates follow-up, home-based care practices as well as the involvement of male caregivers. Resource constraints and negative experiences when seeking follow-up care were the most prominent barriers to completing recommended post-discharge follow up and, thus, represents an area of opportunity to further improve adherence to post-discharge care.

Though the common paradigm of “if sick, seek care” can work well when caregivers can accurately determine when to seek care, during the post-discharge period children may deteriorate quickly, and early signs of such deterioration may be subtle and difficult for parents to identify (22). Caregivers may not recognize overt danger signs of severe illness (23). This lack of recognition is disproportionately the case among the socio-economically disadvantaged who may face additional barriers to effective care seeking (24). Our results clearly demonstrate the positive impact of our discharge education program on caretakers’ perceived knowledge and confidence in providing care to their child during the post-discharge period. As such, the transition of care from facility-based care to home-based care by the children’s caregiver may be more effective if caregivers have the necessary education to easily recognize danger signs and not to delay seeking care when they are present (23). Our results also highlight the importance of caregiver engagement for effective knowledge transfer. This information exchange was largely achieved through consistent, clear, and easy to understand materials (25).

Participants reported the extension of benefits to other family members and to the broader community. Both female and male caregivers who participated in this study reported applying the Smart Discharges principles to their other children and sharing them within their communities. Many participants commented on the economic implications and cost savings when they were able to prevent infections/sepsis and the subsequent hospitalizations/medical care for the whole family. Future work should address how we can strengthen educational engagement concerning the care of clinically vulnerable children at the community level, such as through schools, churches, and other community groups.

Within the formal health system, receptivity to routine follow-up of discharged children is critically important to effective long-term management of sepsis and was a noted barrier to post-discharge care. Education for health workers at local health facilities on post-sepsis syndrome may resolve the confusion that healthcare workers face when a seemingly well child comes for follow-up. Healthcare workers will understand that the primary purpose of the post-discharge follow-up visit is to determine if the child is continuing to make a robust recovery or whether any signs or symptoms are present that warrant further care or investigation. It is critical that the paradigm of sepsis care shifts from a primary focus on acute care to a more holistic focus that does not neglect the critical period of vulnerability following hospital discharge, which has been largely neglected in both policy and practice (5, 24).

When children’s male caregivers were included in the Smart Discharges Program, it fostered their involvement in their child’s care. Often, the phone number used for SMS follow-up was that of the father. This arrangement automatically created engagement with the fathers that otherwise may not have existed. Engagement in care was reflected in an increased interest in the post-discharge care process as well as increased provision for essentials of care, such as food and transportation. Gopal, *et al*., highlight that there is a positive association between male involvement and better maternal and child health outcomes in Uganda (26). They suggest involving male community members, specifically fathers and community leaders who are attuned to the context, to improve the approach to how males are involved in care and to help support sustainable behavior change (26).

Financial constraints were frequently reported as a key barrier to care following discharge, especially with regards to transportation. Many caregivers who participated in this study shared that if they do not have the money necessary for transport, that they will not be able to attend follow up appointments. Financial constraints have been identified as a barrier to follow up care in other health contexts (23) and has prompted a call for further interventions to target patients facing transportation challenges (27). For caregivers with constrained resources, a more cost-effective alternative to improving post-discharge care could have a substantial impact.

When a physical, in-person follow-up is not possible, remote patient assessment via phone or other communication technologies could serve as a viable alternative (28, 29). The COVID-19 pandemic resulted in the expanded use of telemedicine and other technologies to deliver healthcare services in Uganda (30). Such experiences should be leveraged so that healthcare workers and patients/caregiver can further optimize the efficiency and quality of post-discharge care (23). The use of community health workers is widespread in many regions of sub-Saharan Africa, but their work is often not well linked to the broader health system with respect to information transfer. Improved digital linkages between community health workers and their referral facilities could substantially improve the provision of post-discharge care through discharge notifications being sent directly to community health workers (31, 32).

### Limitations

This study is subject to several limitations. First, only children under the age of five were enrolled in the Smart Discharges Program. As such, children themselves did not contribute their perceptions of the program. It is probable that children may have been able to offer a valuable perspective on the Smart Discharges Program, though this may be more pronounced if older children were enrolled into the parent study. Second, the findings of this study were specifically focused on an intervention within the context of a research evaluation. As such, implementation science studies are needed to facilitate integration of this type of program into routine care.

Finally, this study was conducted in a single county and further qualitative work is needed to understand how this approach may be generalizable to other regions.

## Conclusions

The Smart Discharges Program is a digitally assisted health innovation that facilitates discharge follow-up, home-based care practices, and the involvement of male caregivers. Opportunities to improve pediatric discharge care include developing interventions to address caregivers’ resource constraints and negative experiences when seeking follow-up care. The Smart Discharges approach is an impactful strategy to improve pediatric post-discharge care, and similar approaches should be considered to improve the hospital to home transition in other low-resource settings.

## Data Availability

The data generated and analyzed during the current study, which include participant interview and focus group discussion transcripts, are not publicly available due to their potentially directly and indirectly identifiable nature. Furthermore, the open sharing of the qualitative data was not included in the study’s approved protocol nor included in the participant consent forms. Principal investigators can work with interested parties to re-analyze any of the original data if there are queries that are not sufficiently addressed in the manuscript. Reasonable requests can be made to Walimu via corresponding author Dr. Nathan Kenya-Mugisha.

## Acknowledgements

We would like to acknowledge the WALIMU team, staff at the participating hospitals, and patients and caregivers for their participation.

## Author Contributions

This study was conceptualized and designed by MOW in conjunction with NKM, STJ, RS, and OK. MOW and NKM acquired funding for the project. GS, BK, CK, and OK completed data collection and synthesis. OK and JB led the data analysis and interpretation, with initial support from BD and RS. JB led principal drafting of the manuscript, with contributions from OK and support from MOW. All authors critically reviewed and approved the final manuscript.

## References

1. Singer M, Deutschman CS, Seymour CW, Shankar-Hari M, Annane D, Bauer M, et al. The Third International Consensus Definitions for Sepsis and Septic Shock (Sepsis-3). JAMA. 2016;315(8):801–10.

2. Rudd KE, Johnson SC, Agesa KM, Shackelford KA, Tsoi D, Kievlan DR, et al. Global, regional, and national sepsis incidence and mortality, 1990-2017: analysis for the Global Burden of Disease Study. The Lancet. 2020;395(10219):200–11.

3. Akech S, Kwambai T, Wiens MO, Chandna A, Berkley JA, Snow RW. Tackling post-discharge mortality in children living in LMICs to reduce child deaths. The Lancet Child & Adolescent Health. 2023;7(3):149–51.

4. Chaudhry M, Knappett M, Nguyen V, Trawin J, Mugisha NK, Kabakyenga J, et al. Pediatric post-discharge mortality in resource-poor countries: A protocol for an updated systematic review and meta-analysis. PloS one. 2023;18(2):e0281732.

5. Nemetchek B, English L, Kissoon N, Ansermino JM, Moschovis PP, Kabakyenga J, et al. Paediatric postdischarge mortality in developing countries: a systematic review. BMJ Open. 2018;8(12):e023445.

6. Njunge J, Tickell K, Diallo A, Sayeem Bin Shahid A, Gazi M, Saleem A, et al. The Childhood Acute Illness and Nutrition (CHAIN) network nested case-cohort study protocol: a multi-omics approach to understanding mortality among children in sub-Saharan Africa and South Asia [version 2; peer review: 2 approved]. Gates Open Research. 2022;6(77).

7. Wiens MO, Bone JN, Kumbakumba E, Businge S, Tagoola A, Sherine SO, et al. Mortality after hospital discharge among children younger than 5 years admitted with suspected sepsis in Uganda: a prospective, multisite, observational cohort study. The Lancet Child & Adolescent Health. 2023.

8. Wiens MO, Pawluk S, Kissoon N, Kumbakumba E, Ansermino JM, Singer J, et al. Pediatric post-discharge mortality in resource poor countries: a systematic review. PloS one. 2013;8(6):e66698.

9. Ranjeva SL, Warf BC, Schiff SJ. Economic burden of neonatal sepsis in sub-Saharan Africa. BMJ Global Health. 2018;3(1):e000347.

10. Wiens MO, Kumbakumba E, Kissoon N, Ansermino JM, Ndamira A, Larson CP. Pediatric sepsis in the developing world: challenges in defining sepsis and issues in post-discharge mortality. Clinical epidemiology. 2012;4:319–25.

11. Fleischmann-Struzek C, Goldfarb DM, Schlattmann P, Schlapbach LJ, Reinhart K, Kissoon N. The global burden of paediatric and neonatal sepsis: a systematic review. The Lancet Respiratory Medicine. 2018;6(3):223–30.

12. Wiens MO, Kumbakumba E, Larson CP, Moschovis PP, Barigye C, Kabakyenga J, et al. Scheduled Follow-Up Referrals and Simple Prevention Kits Including Counseling to Improve Post-Discharge Outcomes Among Children in Uganda: A Proof-of-Concept Study. Global Health: Science and Practice. 2016;4(3):422–34.

13. Krepiakevich A, Khowaja A, Kabajaasi O, Nemetchek B, Ansermino JM, Kissoon N, et al. Out of pocket costs and time/productivity losses for pediatric sepsis in Uganda: a mixed-methods study. BMC Health Services Research. 2021;21(1):1–9.

14. Wiens MO, Kumbakumba E, Larson CP, Ansermino JM, Singer J, Kissoon N, et al. Postdischarge mortality in children with acute infectious diseases: derivation of postdischarge mortality prediction models. BMJ Open. 2015;5(11):e009449.

15. ClinicalTrials.gov [Internet]. Bethesda (MD): National Library of Medicine (US). 2023 Feb 15. Identifier NCT05730452, Smart Discharges to Improve Post-discharge Health Outcomes in Children; 2023 Feb 15 [cited 2023 May 2]. Available from: https://clinicaltrials.gov/ct2/show/NCT05730452?term=post-discharge+mortality&draw=2&rank=2

16. van Manen M, Higgins I, van der Riet P. A conversation with Max van Manen on phenomenology in its original sense. Nursing & Health Sciences. 2016;18(1):4–7.

17. Sundler AJ, Lindberg E, Nilsson C, Palmér L. Qualitative thematic analysis based on descriptive phenomenology. Nursing Open. 2019;6(3):733–9.

18. Tong A, Sainsbury P, Craig J. Consolidated criteria for reporting qualitative research (COREQ): a 32-item checklist for interviews and focus groups. International Journal for Quality in Health Care. 2007;19(6):349–57.

19. Ministry of Health Uganda. Annual Health Sector Performance Report 2019-2020. *Available from URL* http://www.health.go.ug/mohreports.htm. Published online 2020:222. http://library.health.go.ug/publications/performance-management/annual-health-sector-performance-report-financial-year-201920.

20. Korstjens I, Moser A. Series: Practical guidance to qualitative research. Part 4: Trustworthiness and publishing. European Journal of General Practice. 2018;24(1):120–4.

21. Cohen D. Qualitative research guidelines project. 2006. *Available from* http://www.qualres.org/

22. English L, Kumbakumba E, Larson CP, Kabakyenga J, Singer J, Kissoon N, et al. Pediatric out-of-hospital deaths following hospital discharge: a mixed-methods study. African health sciences. 2016;16(4):883–91.

23. Ranjit S, Kissoon N. Challenges and Solutions in translating sepsis guidelines into practice in resource-limited settings. Translational Pediatrics. 2021;10(10):2646–65.

24. Wiens MO, Kissoon N, Holsti L. Challenges in pediatric post-sepsis care in resource limited settings: a narrative review. Transl Pediatr. 2021;10(10):2666–77.

25. Gourabathini H, Bhalala U. Improving complex discharge process: It’s all about parental education and empowerment. Journal of Pediatric Critical Care. 2022;9(5):155–6.

26. Gopal P, Fisher D, Seruwagi G, Taddese HB. Male involvement in reproductive, maternal, newborn, and child health: evaluating gaps between policy and practice in Uganda. Reproductive Health. 2020;17(1):114.

27. Nkurunziza T, Riviello R, Kateera F, Nihiwacu E, Nkurunziza J, Gruendl M, et al. Enablers and barriers to post-discharge follow-up among women who have undergone a caesarean section: experiences from a prospective cohort in rural Rwanda. BMC Health Services Research. 2022;22(1):733.

28. Christie SA, Mbianyor MA, Dissak-Delon FN, Tanjong MM, Chichom-Mefire A, Dicker RA, et al. Feasibility of a Cellular Telephone Follow-Up Program After Injury in Sub-Saharan Africa. World Journal of Surgery. 2020;44(8):2533–41.

29. Ajayi F. Mobile Communications, physical distance and access to follow-up healthcare service in Lagos Metropolis. Bulletin of Geography. 2018.

30. Louis Henry Kamulegeya JMB, Davis Musinguzi, Pauline Bakibinga. Continuity of health service delivery during the COVID-19 pandemic: the role of digital health technologies in Uganda. The Pan African Medical Journal. 2020;35(2).

31. Feroz A, Jabeen R, Saleem S. Using mobile phones to improve community health workers performance in low-and-middle-income countries. BMC Public Health. 2020;20(1):49.

32. Odendaal W, Lewin S, McKinstry B, Tomlinson M, Jordaan E, Mazinu M, et al. Using a mHealth system to recall and refer existing clients and refer community members with health concerns to primary healthcare facilities in South Africa: a feasibility study. Global Health Action. 2020;13(1):1717410.

